# Parents’ perceptions and intention to vaccinate their children against COVID-19: Results from a cross-sectional national survey in India

**DOI:** 10.1101/2021.10.30.21265449

**Authors:** Bijaya Kumar Padhi, Prakasini Satapathy, Vineeth Rajagopal, Neeti Rustagi, Jatina Vij, Lovely Jain, Venkatesan Chakrapani, Binod Patro, Sitanshu Sekhar Kar, Ritesh Singh, Star Pala, Lalit Sankhe, Bhavesh Modi, Surya Bali, Tanvi Kiran, Kapil Goel, Arun Kumar Aggarwal, Madhu Gupta

## Abstract

**Background:** Despite the success of adult vaccination against COVID-19, providing vaccines to children remains a challenge for policymakers globally. As parents are primary decision-makers for their children, we aimed to assess parents’ perceptions and intentions regarding COVID-19 vaccination in India.

**Methods:** A cross-sectional web-based study was designed, parents or caregivers (N=770) were recruited through snowball sampling using Google form. Cross-tabulation was performed by parents’ intention to vaccinate their children against COVID-19 virus with sociodemographic characteristics and their risk perception towards COVID-19, trust in the healthcare system, and their history of vaccine hesitancy behavior. Multivariable logistic regression analysis was performed to compute the predictors of child vaccination intention among Indian parents.

**Results:** 770 parents across the country have completed the survey. Of the 770 participants, 258 (33.5%) have shown intent to vaccinate their children. The stated likelihood of child vaccination was greater among parents who had a bachelor’s degree or higher education (aOR: 1.98, 95% CI: 1.15-3.51); as well as among parents who intended to vaccinate themselves (aOR: 2.35, 95% CI: 1.30-4.67). Parental concerns centered around vaccine safety and side effects.

**Conclusion:** Indian parents reported high knowledge of the COVID-19 virus and were aware of the development of a novel vaccine. However, about one-third of parents intended to vaccinate their children, and about half of them were not sure whether to vaccinate their children or not against the COVID-19 virus. The study highlighted the need for health promotion strategies that promote vaccine uptake among parents.

## Introduction

The World Health Organization (WHO) has reported more than 242 million confirmed cases of COVID-19 worldwide, including approximately 5 million deaths as of 20 October 2021(1). India reported 34 million confirmed cases and 450 thousand deaths as of 25^th^ October 2021(1). An effective strategy to mitigate the morbidity and mortality of the COVID-19 and to ensure population higher levels of immunity is the development of an effective and safe vaccine for all populations, including children. In general, children with COVID-19 present with milder symptoms and are at lower risk of hospitalization and life-threatening complications(2). Numerous public health authorities, including the WHO and United States Centers for Disease Control and Prevention, advocate a vaccine for children. However, acceptance of the COVID-19 vaccine among parents or caregivers remains unexplored.

Since the start of the COVID-19 pandemic, a high number of vaccines is being developed(3). The WHO has declared on the 11^th^ of July 2021 that 107 and 184 vaccine candidates are in clinical and pre-clinical stages, respectively, and at least 13 vaccines have been approved and administrated through four platforms(4). As of 25 October 2021, approximately 6 billion vaccine doses have been administrated worldwide(1). In India, as of 25 October 2021, the Ministry of Health and Family Welfare reported over one billion doses to adults(5). To expand the vaccine administration to children, the government of India is planning to implement a COVID-19 vaccine campaign for the age group of 12 to 18 years old(6).

Numerous studies on the COVID-19 vaccine acceptance shows that a lower number of parents were interested in vaccinating their children(7–14). A national survey in the United States reported less than one-half of parents are likely to administer a COVID-19 vaccine to their children(15). In Canada, 63% of parents intend to vaccinate their children against COVID-19(10). In Turkey, only 36.3% of parents were willing to have their children receive the COVID-19 vaccine(13). In China, COVID-19 vaccine hesitancy was found to be 8.44% among reproductive women(8). In a hospital-based study in Saudi Arabia, one-fourth of mothers were hesitant towards childhood immunization(11). To the best of our knowledge, none of the studies conducted in India address the intention and perception of parents’ willingness to vaccinate their children in India.

Despite the success of adult vaccination globally, childhood immunization against the COVID-19 vaccine is now a challenge for policymakers. Including an effective and safe vaccine, a high level of vaccine acceptance among the parents or caregivers is also a key factor for global immunization success. Many factors such as fear about the vaccine safety and risk for a child to be infected by the virus impact parents’ acceptance of the new COVID-19 vaccination program(13). Therefore, understanding parents’ perception and acceptance of the COVID-19 vaccine will help in designing effective strategies for promotion and rolling out COVID-19 vaccination and beyond. To the best of our knowledge, this is the first community-based study in India that investigates parental willingness for COVID-19 vaccination. The study also explored factors associated with vaccine hesitancy among Indian parents or caregivers.

## Methods

### Study Design and Sample

We conducted a national online cross-sectional survey between November 2020 and January 2021 (before the introduction of the novel COVID-19 vaccine). The overall sample (N = 770) of parents were recruited across major geographical regions in India with an equal weightage to the urban and rural population. Respondents were adults who have one or more children 0–17 years old in their home and have access to the internet or telephone. We estimated a minimal sample size of 768, based on the maximum variability possible in the outcome variable in the population (i.e., a proportion of 0.50), with a margin of error of +/-5% and 95% confidence intervals (CI). The design effect was kept at 2. Study participants were recruited through snowball sampling. Invitations to participate in the study were distributed to the respondents via email, social media: Twitter, Facebook, and the WhatsApp communication platform of the primary contacts of the study team and requested to transmit further for its maximum reach. Considerations were made to recruit the participants across major geographic regions of India. To ensure rigor and validity, respondents had unique IDs, and 10% of respondents were contacted by telephone for cross verification of the presence of children 0–17 years old in their home.

### Questionnaire Development

A first draft of the questionnaire was prepared based on a review of the relevant literature and reviewed by subject experts. The draft questionnaires were pre-tested and then pilot tested with 50 participants and adjusted for accuracy and clarity. The consistency and stability of the final questionnaires were tested using Cronbach’s alpha (0.7). The online survey was administered via Google Forms and was distributed to the parents via Facebook, WhatsApp, and mail groups. Informed consent was obtained from all parents (18 years or older and currently living in India). Detailed responses were recorded further only those who provided informed consent to complete the survey.

#### Sociodemographic Measures

The survey has detailed information about the sociodemographic characteristics of study participants, including parental age, sex, education level, place of residence, income, occupation, social status in the community.

#### Parents knowledge of COVID-19 and vaccine intention

The survey tested parents’ knowledge of the COVID-19 virus by asking, “Before this interview, were you aware that the COVID-19 virus is currently circulating in the community?”. The responses were captured in a 3-point Likert scale (Yes, No, and Don’t know). We also asked parents to record their response to vaccine development knowledge by asking, “To the best of your knowledge, is there currently a vaccine being prepared for the pandemic Coronavirus strain referred to as COVID-19 vaccine?”. Parents’ own likelihood of getting a COVID-19 vaccine (same 3-point Likert scale: Yes, No, and Not sure) was also recorded.

#### Intent to vaccinate the child

The survey asked, “Do you intend to vaccinate your child(ren) for COVID-19 once a vaccine is available for children?”. The responses were captured in a 3-point Likert scale (Yes, No, and Not sure). We coded “Yes” as “likely to get a COVID-19 vaccine”; all others were labeled as “hesitant.”

### Statistical Analyses

The key outcome measures of the study were to know the parent’s intention to vaccinate their children against the COVID-19 virus. Those who responded “Yes” were labeled as “Intended to vaccinate” vs. “hesitancy.” Descriptive statistical analysis was performed by doing cross-tabulation of demographic characteristics with the primary response variable “Intended to vaccinate.” We performed both simple and multivariable logistic regression analyses to compute the odds ratio (OR) and a 95% confidence interval (CI). Chi-squared tests were performed for bivariate analysis (cross-tabulation) between the outcome variable and all explanatory variables. A multivariate logistic regression model was constructed by including factors with P-values <.05 at binary comparisons. Inference on significant association was considered with a two-tailed p≤0.05. STATA 15.0 software (StataCorp LP, Texas, USA) was used for all statistical analyses.

### Ethical Considerations

Ethical approval was granted for the study by the institutional Research Ethics Committee of Post Graduate Institute of Medical Education and Research(PGIMER), Chandigarh, India. Informed assent was taken before participation in the study. Anonymized data was used for analysis, interpretation, and reporting.

## Results

Seven hundred and seventy parents participated in this study. Over half of the parents were 30-49 years old (60.0%), 39.6% were female, 28.5% were educated to graduate level or higher, 23.4% employed in government sector, 59.7% had more than six members in their family, 31.8% had a monthly income above 50,000 Indian rupees, 41.3% reside in rural areas, and 48.2% perceived their social status as high in the community (Table 1).

**Table 1:** Sociodemographic characteristics of the study respondents (N=770)

| **Variables** | **n (%)** |
| --- | --- |
| **Parental age** |  |
| 18-29 | 120 (15.6) |
| 30-49 | 462 (60.0) |
| above 50 | 188 (24.4) |
| **Parental sex** |  |
| Male | 465 (60.4) |
| Female | 305 (39.6) |
| **Parental education** |  |
| No Formal Education | 26 (3.4) |
| Primary School | 193 (25.1) |
| Diploma/High School | 262 (34.0) |
| Undergraduate | 130 (16.9) |
| Postgraduate and above | 89 (11.6) |
| **Parent's employment status** |  |
| Government sector | 180 (23.4) |
| Private sector | 282 (36.6) |
| Self-employed | 230 (29.9) |
| Unemployed | 78 (10.1) |
| **Family size** |  |
| Five and below | 310 (40.3) |
| Six and above | 460 (59.7) |
| **Family income (monthly, INR)** |  |
| below 10000 | 52 (6.8) |
| 11000-20000 | 291 (37.8) |
| 21000-50000 | 182 (23.6) |
| above 50000 | 245 (31.8) |
| **Social status** |  |
| Low | 108 (14.0) |
| Medium | 291 (37.8) |
| High | 371 (48.2) |
| **Region of residence** |  |
| Eastern | 192 (24.9) |
| Western | 91 (11.8) |
| Northern | 210 (27.3) |
| Southern | 108 (14.0) |
| Central | 86 (11.2) |
| North-east | 83 (10.8) |
| **Area of residence** |  |
| Urban | 452 (58.7) |
| Rural | 318 (41.3) |
| **Social caste** |  |
| Other Backward Caste | 312 (40.5) |
| Unreserved | 215 (27.9) |
| Scheduled Caste | 150 (19.5) |
| Scheduled Tribe | 93 (12.1) |
| **Religion** |  |
| Hindu | 367 (47.7) |
| Muslim | 173 (22.5) |
| Christian | 85 (11.0) |
| Sikhs | 105 (13.6) |
| Other | 40 (5.2) |

The rate of parents’ willingness to allow their children to receive the COVID-19 vaccine was 33.5%, whereas 40.3% of parents were willing to receive the COVID-19 vaccine. At the time of survey, 27.5% of parents had a history of exposures to a confirmed COVID-19 cases and 62.7% had concerned of getting infected to COVID-19 virus. Most (88.3%) of the parents were aware that COVID-19 virus is currently circulating in the community, and 86.0% of them had knew that a vaccine is being prepared for the pandemic Coronavirus strain referred to as “COVID-19 vaccine”. Of the 770 parents, about one-fourth of them had a previous history of vaccine hesitancy, 62.2% had trusted the healthcare system, and 39.1% considered domestic vaccines are good (Table 2).

**Table 2:** Willingness to accept COVID-19 vaccine, contact history with COVID-19 patients, risk perception, and vaccine preferences among parents (N=770)

| **Variables** | **n(%)** |
| --- | --- |
| Intended to vaccinate the child |  |
| Yes | 258 (33.5) |
| No | 164 (21.3) |
| Not sure | 348 (45.2) |
| Intended to vaccinate themselves |  |
| Yes | 310 (40.3) |
| No | 270 (35.1) |
| Not sure | 190 (24.7) |
| Exposed to COVID-19 cases |  |
| No | 212 (27.5) |
| Yes | 558 (72.5) |
| Knowledge about the COVID-19 virus |  |
| No | 90 (11.7) |
| Yes | 680 (88.3) |
| Knowledge about the development of the COVID19 vaccine |  |
| No | 108 (14.0) |
| Yes | 662 (86.0) |
| History of vaccine hesitancy |  |
| Yes | 210 (27.3) |
| No | 560 (72.7) |
| High-risk perception |  |
| Yes | 483 (62.7) |
| No | 287 (37.3) |
| Trust in the healthcare system |  |
| No | 291 (37.8) |
| Yes | 479 (62.2) |
| Trust in domestic vaccines |  |
| Better | 301 (39.1) |
| Neutral | 282 (36.6) |
| Worse/Not Sure | 187 (24.3) |

Table 3 shows the analysis of bivariate and multivariable logistic models, and determined the predictors affecting parents’ willingness to allow their children to be given the COVID-19 vaccine. In bivariate model, trust in the healthcare system, higher risk perception, domestic vaccine, female respondent, employed in government sector, and being a rural residents were found to be associated with parent’s intention to vaccinate their children. After adjusting for confounding variables, parent’s own willingness to receive the COVID-19 vaccine (aOR: 2.35, 95% CI: 1.30-4.67), and parent’s who had a bachelor’s degree or higher education (aOR: 1.98, 95% CI: 1.15-3.51) were found to be associated with parents’ willingness to receive the COVID-19 vaccine for their children (Table 3).

**Table 3:** Unadjusted and adjusted odds ratios for the association between parent’s intention to vaccinate their children with sociodemographic and other COVID-19 behaviors (N=770)

|  | "Intended to vaccinate children" |  |  |
| --- | --- | --- | --- |
| Variable | OR [95% CI] | aOR [95% CI] | P-value |
| **Parents' COVID-19 vaccination intention for themselves** |  |  |  |
| No | Ref | Ref |  |
| Yes | 2.88 [1.21-4.06] | 2.35 [1.30-4.67] | 0.001 |
| **Exposed to COVID-19 cases** |  |  |  |
| No | Ref | Ref |  |
| Yes | 1.71[0.81-2.61] | 1.28 [0.46-2.25] | 0.311 |
| **Trust in the healthcare system** |  |  |  |
| No | Ref | Ref |  |
| Yes | 2.42 [1.41-3.55] | 2.11 [0.98-3.17] | 0.068 |
| **History of vaccine hesitancy** |  |  |  |
| Yes | Ref | Ref |  |
| No | 1.48 [0.81-3.66] | 1.45 [0.27- 2.74] | 0.241 |
| **Higher risk perception** |  |  |  |
| No | Ref | Ref |  |
| Yes | 2.46 [1.19-3.80] | 1.87 [0.99-3.17] | 0.067 |
| **Opinion in domestic vaccines** |  |  |  |
| Worse | Ref | Ref |  |
| Better | 2.72 [1.02-5.08] | 1.67 [0.71-3.97] | 0.165 |
| Neutral | 2.30 [1.04-2.87] | 1.40 [0.26-2.83] | 0.151 |
| **Age** |  |  |  |
| 18 – 29 | Ref | Ref |  |
| 30-49 | 1.93 [0.85-2.23] | 1.13 [0.48-2.77] | 0.126 |
| above 50 | 1.78 [0.91-2.14] | 1.52 [0.35-2.17] | 0.132 |
| **Sex** |  |  |  |
| Male | Ref | Ref |  |
| Female | 1.97 [1.07-2.20] | 1.35 [0.97-2.44] | 0.081 |
| **Employment status** |  |  |  |
| Unemployed/self-employed | Ref | Ref |  |
| Private sector | 1.51 [0.65-2.20] | 1.18 [0.31-2.70] | 0.142 |
| Government sector | 1.91 [1.07- 3.31] | 1.30 [0.71-2.67] | 0.095 |
| **Education** |  |  |  |
| Primary or below | Ref | Ref |  |
| Diploma/High School | 2.18 [1.19-5.82] | 1.86 [0.95-4.61] | 0.087 |
| Undergraduate and higher | 3.41[1.32-6.11] | 1.98 [1.15-3.51] | 0.001 |
| **Social status in the community** |  |  |  |
| Low | Ref | Ref |  |
| Medium | 1.80 [0.59-2.72] | 1.57 [0.92-2.67] | 0.085 |
| High | 1.83 [0.58-2.88] | 1.16 [0.75-2.24] | 0.116 |
| **Place of residence** |  |  |  |
| Urban | Ref | Ref |  |
| Rural | 1.92 [1.07-3.12] | 1.25 [0.44-3.05] | 0.134 |

Figure 1, shows the concerns of parents’ in accepting the novel COVID-19 vaccine. The key reasons for reluctance to allow their children to receive the COVID-19 vaccine included safety and effectiveness (86.4%), side effects (78.2%), doses not known (63.7%), lack of sufficient scientific data (53.2%), childrens are not affected from COVID-19 (52.8%), and trust in vaccine (43.4%).

**Figure 1:**
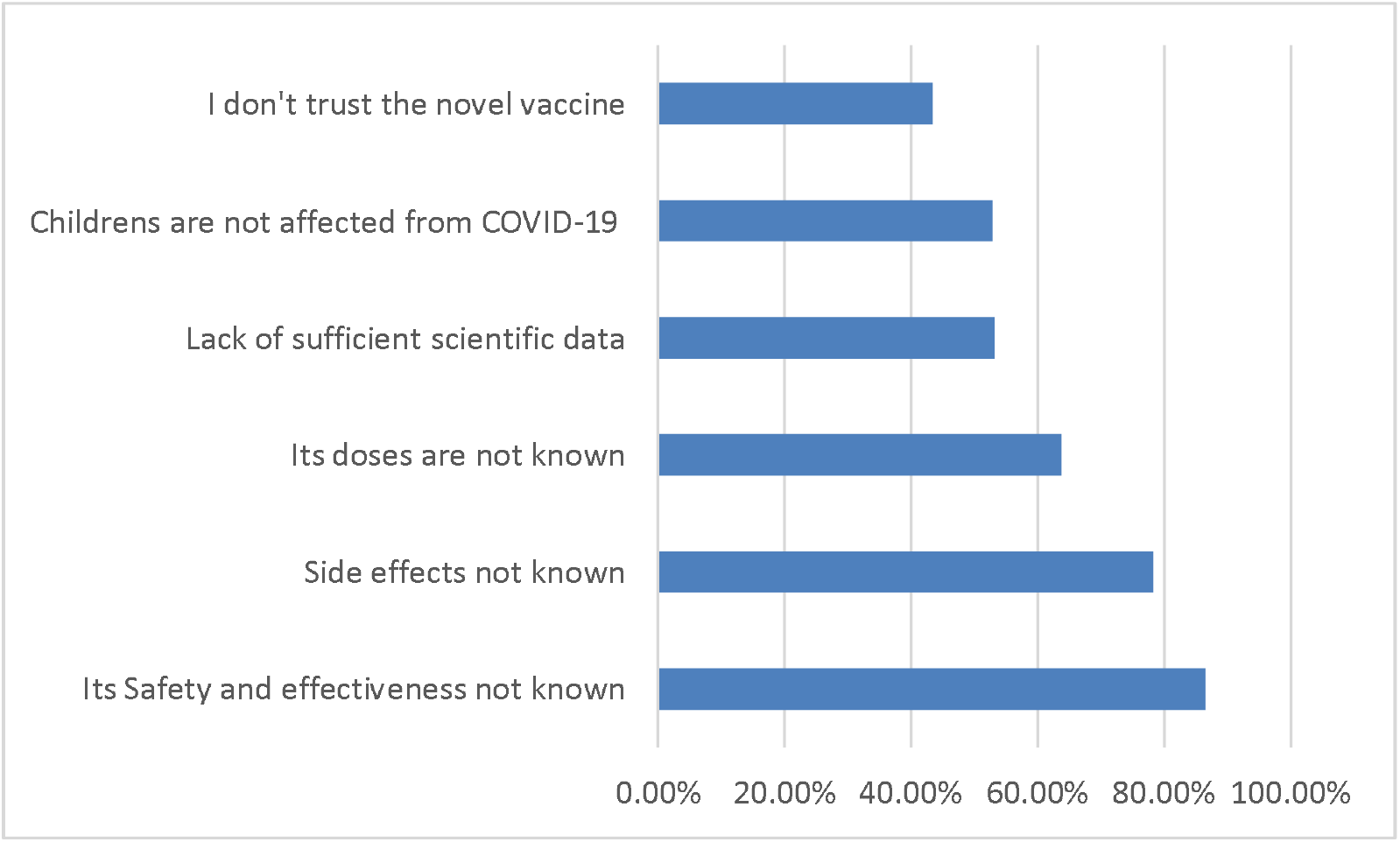
Parents’ concerns for not willing to vaccinate of their children for COVID-19 (n = 770)

## Discussion

In this study, we aimed to determine the acceptability of the COVID-19 vaccine among parents in India and the associated factors. To the best of our knowledge, this is the first study in India that investigates parents’ willingness to administer COVID-19 vaccine to their children. Understanding the factors associated with the intention will help in implementation of vaccination program in India and elsewhere. We found that nearly 33.5% of parents in India would be willing to administer a COVID-19 vaccine to their child. Sociodemographic factors, such as higher education level and parents who intended to vaccinate themselves, were found to be associated with childhood vaccination. The main concerns raised by parents in our study were around the safety and effectiveness of the novel COVID-19 vaccine.

Our findings contribute to an emerging literature suggesting that there is a low demand for childhood COVID-19 vaccine among parents(7–9,11,16). A study conducted in Texas, USA reported that about 38.3% of mothers had no intention to consider COVID-19 vaccine for their children(16). Proportion of willing parents in India is considerably lower as compared to other countries as per studies from England (89%)(7); New Zealand (80%)(17); China (73%)(18); USA (65%)(19); Canada (63%)(10); and Turkey (42%)(20). A large proportion of participants in our study were aware about COVID 19 virus(88.3%) ; developmental status of COVID 19 vaccine (86%) and were exposed to COVID 19 (72.5%). Inspite of that, less than half of partcipants were willing to get themselves readily vaccinated (40.3%). This supports the findings that parent’s intention to themselves get vaccinated is the most influential factor determining their intent towards children vaccination. Factors such as newness of vaccine, rapid development and unknown long term side effects influence parent’s perception of vaccine safety and their intention to get themselves and their children vaccinated(13). As reported by numerous studies, vaccine safety is fundamental to maintaining the public’s trust in vaccines(12,21,22). Thus future studies are required to understand long term change in perception of parents and factors influencing their intention to get themselves and their children vaccinated. Health care providers play a key role in influencing decision of parents towards vaccine acceptance and uptake. It is essential to thus engage various level of health care providers and strategize vaccine messages to parents especially from lower education status

Limitations of this study that should be considered include the sampling strategy adopted by the study which may not be representative of all parents in the country, hence limits the generalizability of our finding. The cross-sectional design of the study should be interpretated carefully when accessing overall prevalence of vaccine hesitancy among parents in India. Our sample represented individuals with internet access and having mobile literacy. This does not necessarily reflect the country wide perception of parents towards vaccine and their intention to get children vaccinated.

Despite the above limitations, our study assessed nationally representative sample of parents regarding their perceptions and intentions to vaccinate their children at a critical time, just as adult COVID-19 vaccination programs was initiated in India. Our findings reminiscent of recent research suggesting that there is a low demand for COVID-19 vaccine among parents, and highlights the need for longitudinal studies to measure the acceptability of a COVID-19 vaccine at different intervals. Future studies are thus required to supplement our current findings to enhance vaccine uptake among children in India. Nevertheless, our study establishes evidence regarding parent’s hesitancy towards getting their children vaccinated and their concerns in coming months must be addressed through targeted communication stratgeies.

## Conclusion

This study depicts that Indian parents have good knowledge of COVID-19 and vaccine developments. However, the demand for COVID-19 vaccine among parents was found to be low, with many parents unsure about their acceptance of COVID-19. The study warents implementation of strategies to improve parents’ knowledge about the safety, efficacy and side effects of COVID-19 vaccine.

## Data Availability

All data produced in the present study are available upon reasonable request to the authors

## Abbreviations

COVID-19: Coronavirus disease-2019
WHO: World Health organization

## Acknowledgements

The authors would like to acknowledge the study participants for their time and contributions to the study.

## Authors’ contributions

BKP, MG and AKG conceptualized the study, designed the tools; BKP, LJ, JV, PS, VC, BP, SSK, RS, SP, SB, NR, VR, TK, KG, BM, LS, MG, AKG conducted the study at national level, and collected the data. All authors reviewed drafts, provided edits, and approved the final submission.

## Availability of data and materials

De-identified data is available upon request.

## Declarations

### Ethics approval and consent to participate

The study protocol was reviewed and approved by the Institutional Ethical Committee of PostGraduate Institute of Medical Education and Research (PGIMER), Chandigarh, India. Informed consent was obtained from all participants. Anonymized data was used for interpretation and reporting.

### Consent for publication

Not applicable.

